# Incidence Rate of Post Coronary Artery Shunt Complications; Age Dependent!

**DOI:** 10.1101/2022.12.26.22283945

**Authors:** Basheer Abdullah Marzoog, Ekaterina Vanichkina

## Abstract

**Background:** Several complications reported after coronary artery bypass graft surgery (CABG), such as postoperative arrythmia and postoperative stroke.

**Aims:** The study aimed to assess the role of age advancement in the incidence rate of post coronary artery shunt complications.

**Objectives:** Aging is a pathophysiological process that experience every living cell and starts from the early period of development, before even born.

**Materials and methods:** A retrospective analysis of 290 patients who underwent a coronary artery bypass graft at Mordovia republic hospital for the period 2017-2021. The sample was subdivided into two groups, the first group 126 patients (mean age: min; max, 55.21:41.00;60.00) and the second group 163 (M age: min; max, 66.11: 61.00; 80.00). For statistical analysis, we used the T test, the one-way ANOVA test, ROC analysis, and the Pearson correlation test using the Statistica 12 programme.

**Results:** Post-CABG arrythmia developed in elderly patients, p<0.012528. Subsequently, after CABG arrythmia, the ICU and total hospitalisation days, p< 0.000000; p< 0.000072, respectively. Elderly people are at higher risk of psychosis after CABG surgery, p <0.007379. psychosis significantly increases hospitalisation days in the ICU, p < 0.000140. Postoperative stroke occurs more frequently in elderly people, p < 0.037736. Subsequently, postoperative stroke increases the ICU hospitalization days, p <0.000747. The number of ITAs used is less in the elderly, p < 0.016145. In terms of correlation, exist a direct association between age and ICU / total hospitalization days/number of complications, r= 0.189046, 0.141415, 0.138565; respectively.

**Conclusions:** The number of complications is determined by age, CPB time, aortic cross-clamp time, the days of hospitalisation in the ICU and the total days of hospitalisation. Elderly people undergoing CABG are at higher risk of psychosis, arrythmia, longer total and ICU hospitalization days, and stroke.

**Others:** Total hospitalization days depend on the presence of arrythmia, which is commonly seen in elderly patients > 63 years old.

**Graphical abstract:** Graphical abstract:
With age, the number of shunts increases in the first group <60 years old.

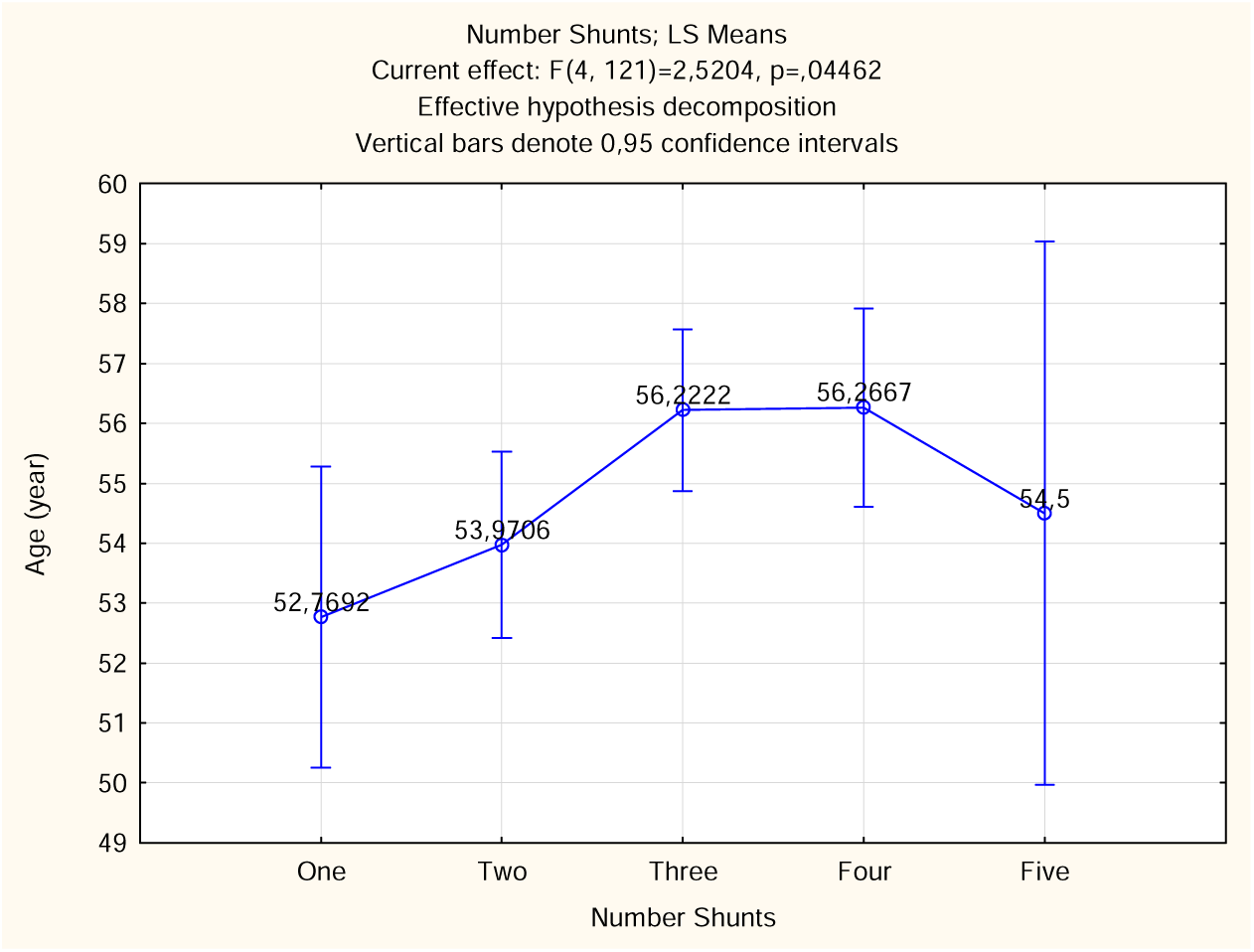

## Background

Coronary artery graft surgery (CABG) performed on patients with ischemic heart disease to relieve the discrepancy between demand and supply of heart tissue. CABG surgery is performed in two versions, with stopped heart (on-pump) or working heart (off-pump) with cardiopulmonary bypass (CPB). CABG surgery involves revascularization of the coronary arteries through grafting of vessels from the saphenous vein, brachial vein, internal thoracic arteries, internal mammary arteries, and intercostal thoracic arteries [1]. The surgical approach to CABG is performed endoscopically or by thoracotomy. Recent surgeries are performed using the Da-Venice robot to minimize the risk of complications and improve prognosis by reducing the recovery period [2,3].

Post CABG complications are frequent and seen in all types of CABG surgery, off-pump and on-pump. The remarkable advancement in the current CABG techniques reduced the complications frequency, but the underlying risk factors and pathophysiological mechanisms of development of each complication remain unclear. Depending on the type of surgery, the risk of development of complication is different. Several complications are reported after the postoperative period, such as postoperative arrythmia, post-operative psychosis, postoperative stroke, postoperative myocardial infarction, postoperative hydrothorax, re-sternotomy, postoperative dyspnea, and sternal wound infection [3,4]. The risk factors for development of post CABG include diabetes mellites, arterial hypertension, dyspnea, progressive angina, and post myocardial infarction (MI) sclerosis, advanced age, long CPB time, prolonged Aortic cross-clamp time, long ICU hospitalization days, and long total hospitalization days [5,6].

## Materials and methods

A retrospective analysis of 290 patients who underwent a coronary artery bypass graft at Mordovia republic hospital for the period 2017-2021. The sample was divided by age into two groups A (60≥age in years ≥41) and B (80≥age in years ≥61). Accordingly, we used the stepwise model builder-logistic regression. The used model variables in the test are ejection fraction, age in numbers, BMI, CPB, XCL time, days reanimation, total days after surgery, and number of complications. We found that age plays important role. (*Figure 1*) (*Table 1*)

**Figure 1:**
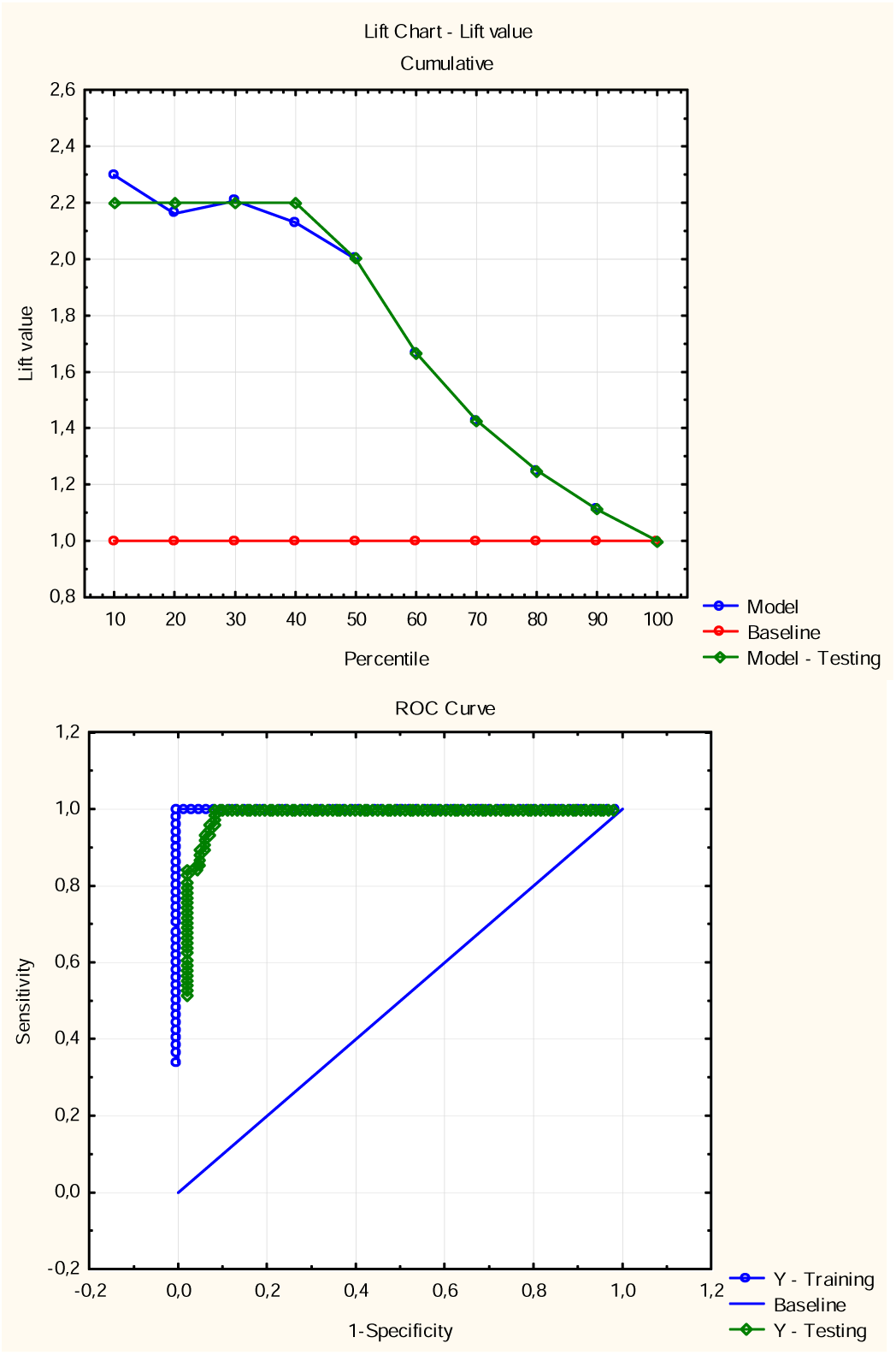
Receiver Operating Characteristic (ROC) Curve Analysis and Area Under the curve (AUC) analysis of the Sample.

**Table 1:** Predicted and real values of the sample divided by age.

|  |  |  |  |  |  |  |
| --- | --- | --- | --- | --- | --- | --- |
|  | Classification of cases (stat in copy of post coronary artery shunt complications)Odds ratio: infinityLog odds ratio: infinity |  |  |  |  |  |
|  | Predicted: A | Predicted: B | Percent correct | Predicted: A - Testing | Predicted: B - Testing | Percent correct - Testing |

|  | Classification of cases (stat in copy of post coronary artery shunt complications)Odds ratio: infinityLog odds ratio: infinity |  |  |  |  |  |
| --- | --- | --- | --- | --- | --- | --- |
|  | Predicted: A | Predicted: B | Percent correct | Predicted: A - Testing | Predicted: B - Testing | Percent correct - Testing |
| Observed: A | 50 | 0 | 100 | 69 | 5 | 93.243 |
| Observed: B | 0 | 61 | 100 | 7 | 91 | 92.857 |

Therefore, the sample has been divided by age into two groups, the first group included 126 patients (mean age: min; max, 55.21:41.00;60.00) and the second group 163 (mean age: min; max, 66.11: 61.00; 80.00). The data were collected from the archive of the hospital for the period 2017-2021. Retrospective analysis has been carried out to assess the role of age advancement in postoperative complications. The mean age of the sample by year is presented in Figure 2. The missing data in different variables has not been included in the absolute values but included in the total percentage of the presented relative values if exist. Assess the hazard ratio using the special tests because of the inability to follow the patients. Also, we couldn’t asses the survival rate due to the same reason.

**Figure 2:**
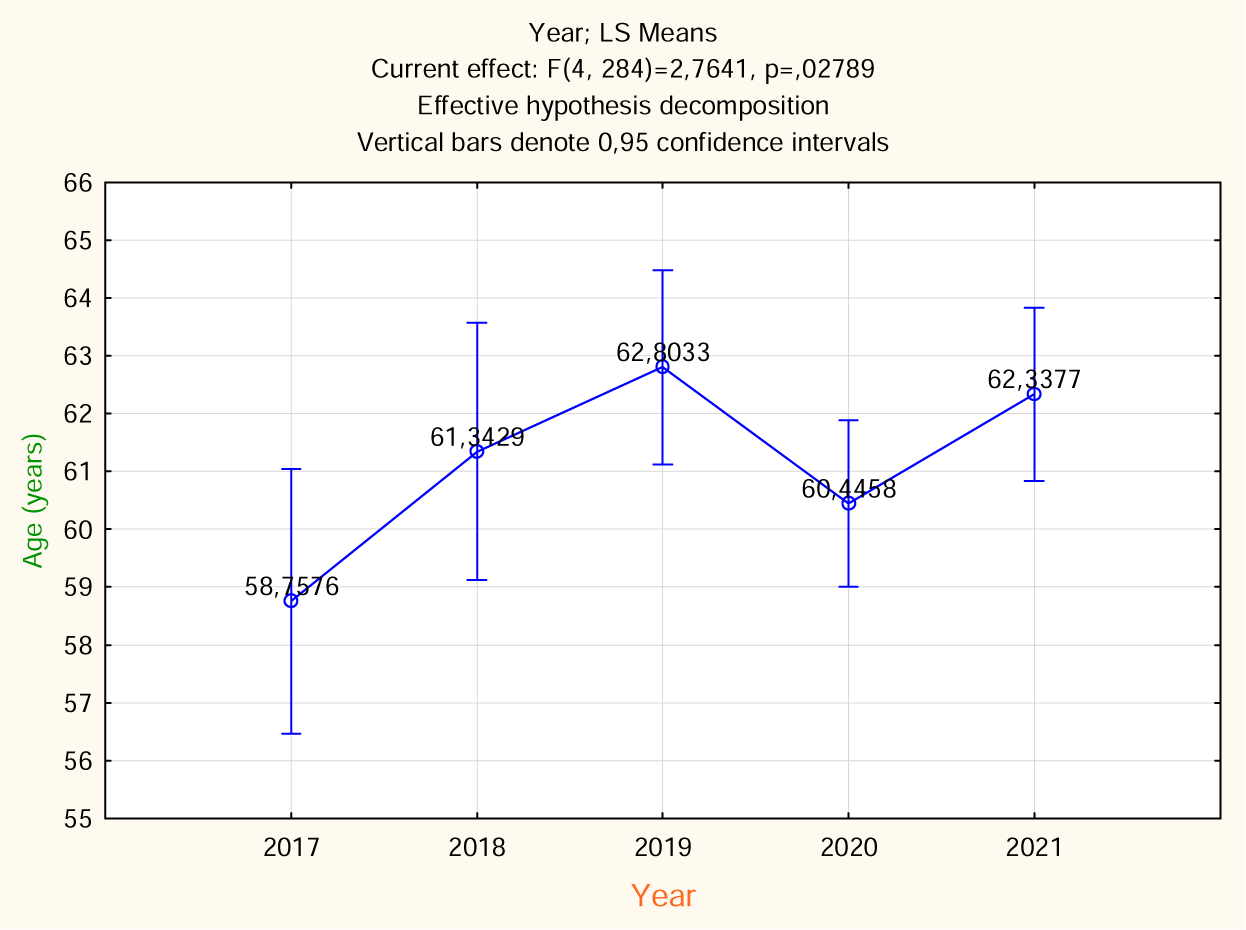
Mean age of the patients who passed coronary artery shunt surgery (whole sample). At the same time this mean age shows average age of occurrence of indications for CABG, which is usually associated with the acute coronary syndrome development.

For statistical analysis, we used the T test, the one-way ANOVA test, and the Pearson correlation test using the Statistica 12 programme. (StatSoft, Inc. (2014). STATISTICA (data analysis software system), version 12. www.statsoft.com.)

## Results

In 2017, 33 (11.37931 %) patients performed coronary bypass surgery. In 2018, the number increased to 35 (12.06897 %) patients. In 2019, the number significantly increased to 61 (21.03448%) patients. In 2020, the highest number of bypass surgeries performed, 84 (28.96552 %) patients. In 2021, 77 (26.55172%) patients underwent coronary bypass surgery. Body mass index (BMI) range from 16.360-45.350 Kg/m^2^ (mean; standard error: 45.350; 0.241918). Ejection fraction ranged from 29.000-77.000 % (mean; standard error: 55.772; 0.438403). 278 (95.86207%) patients underwent on-pump CABG surgery and 12 (4.13793 %) patients underwent off-pump CABG surgery. The period of artificial circulation in CPB surgeries ranged from 0.000-431.000 minute (M; ±m: 431.000; 2.661081). The ischemia time ranged from 0.000-147.000 minute (M; ±m: 55.466; 1.341570). Postoperative hospitalisation (hereafter, postoperative and post-CABG will be used interchangeably) in the intensive care unit ranged from 1.000 to 27.000 days (M; ±m: 2.821; 0.140122). The total hospitalization days (ICU hospitalization days plus recovery hospitalization days) after surgery ranged 7.000-50.000 days (M; ±m: 11.722; 0.292762).

In the first group, the mean age is 55.21 years (min; max, 41.00; 60.00) and the mean BMI is 28.8208 kg/m2 (min; max, 16.36; 45.35). Moreover, in the first group, mean ejection fraction is 54.85% (min; max, 29.00; 76.00) and the mean CPB 101.19 minutes (min; max, 0.00; 270.00). Furthermore, the mean XCL time is 56.29 minute (min; max, 0.00; 105.00) and the mean hospitalisation days in the intensive care unit is 2.4758 days (min; max, 1.00; 6.00). Furthermore, the mean total hospitalization days after surgery is 10.96 days (min; max, 7.00000; 26.0000) and the mean number of complications is 0.2857 (min; max, 0.00; 3.00). Non-parametric parameters have been evaluated in table 2. With age, the number of shunts increases in the first group. However, a reduction in the incidence rate of complications has been observed in the second group (*Figure 3)*.

**Figure 3:**
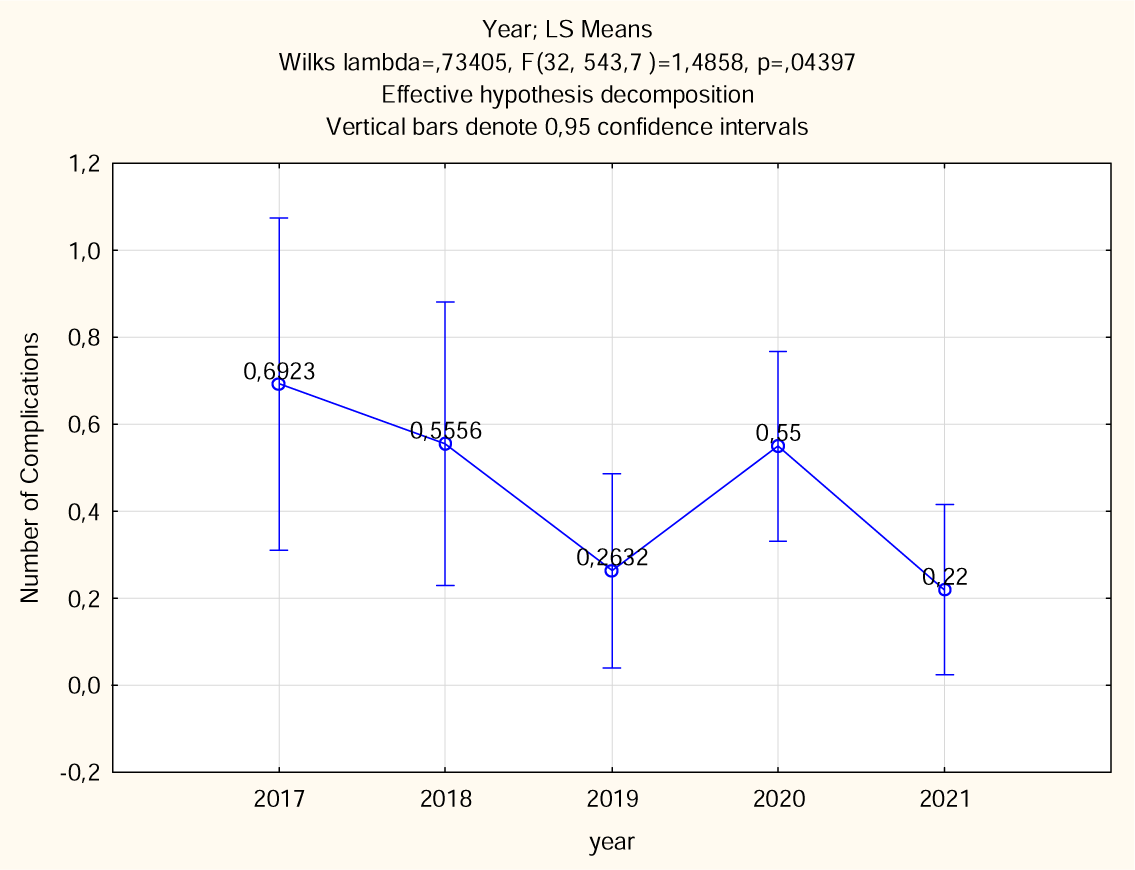
Reduction in the incidence rate of complications in the second group > 60 years.

**Table 2:**
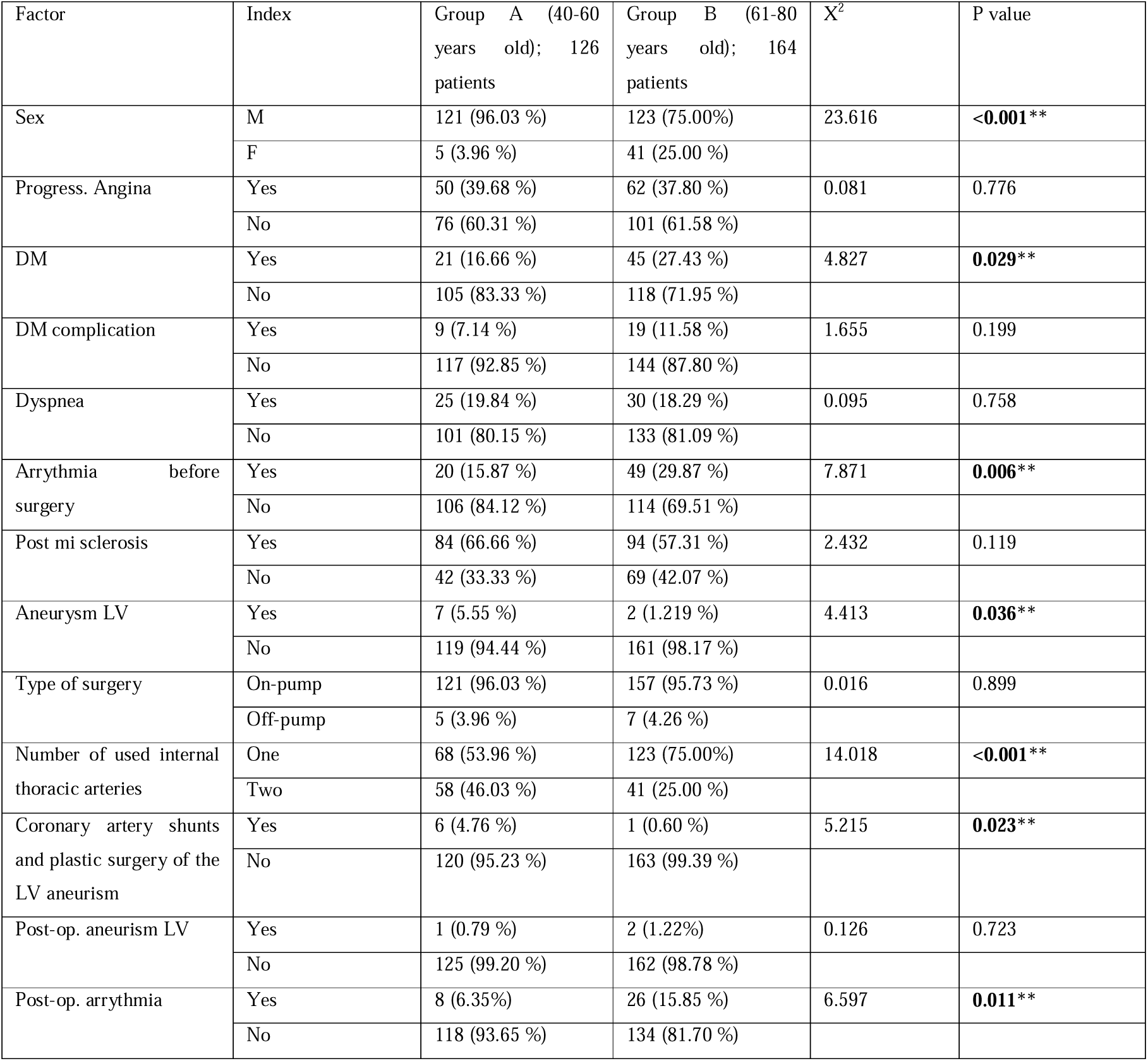
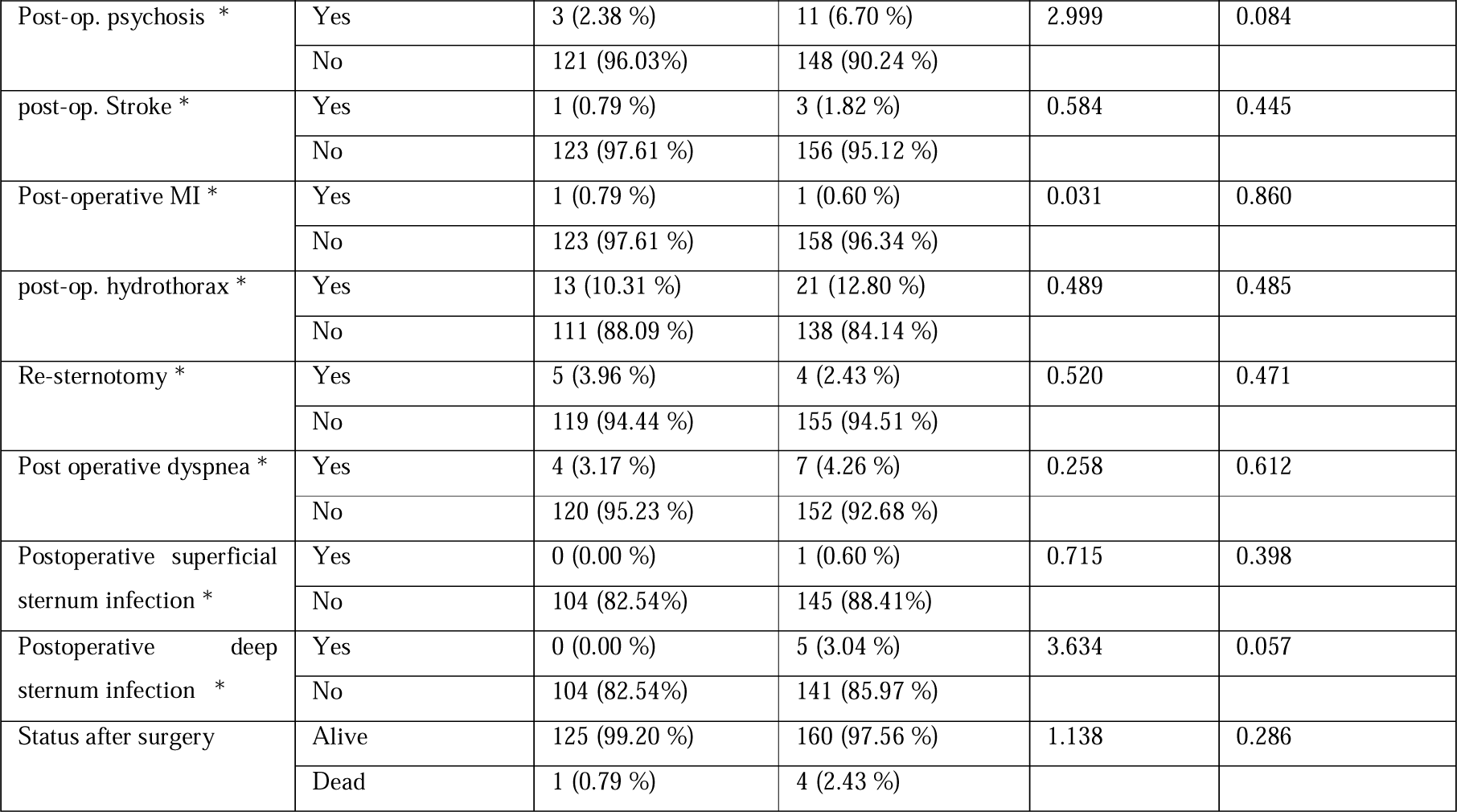
Age in the determination of preoperative conditions and post coronary artery shunt surgery complications. *The missed data are treated as part of the of the relative value but not the absolute value. ** values in bold are statically significant.

In table 3, a comparative demonstration of various factors in the development of different complications in pre- and post- CABG survivals.

**Table 3:**
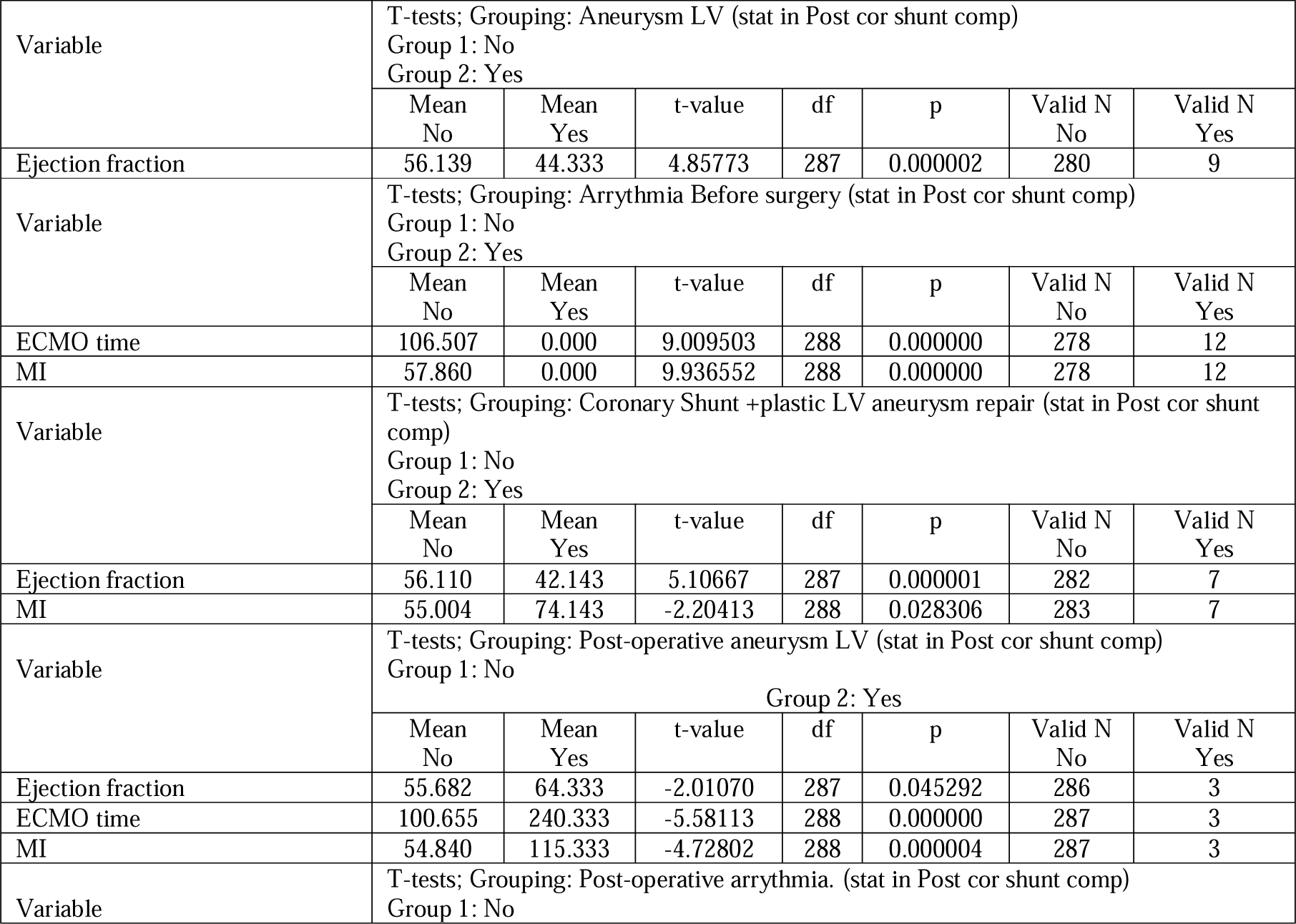
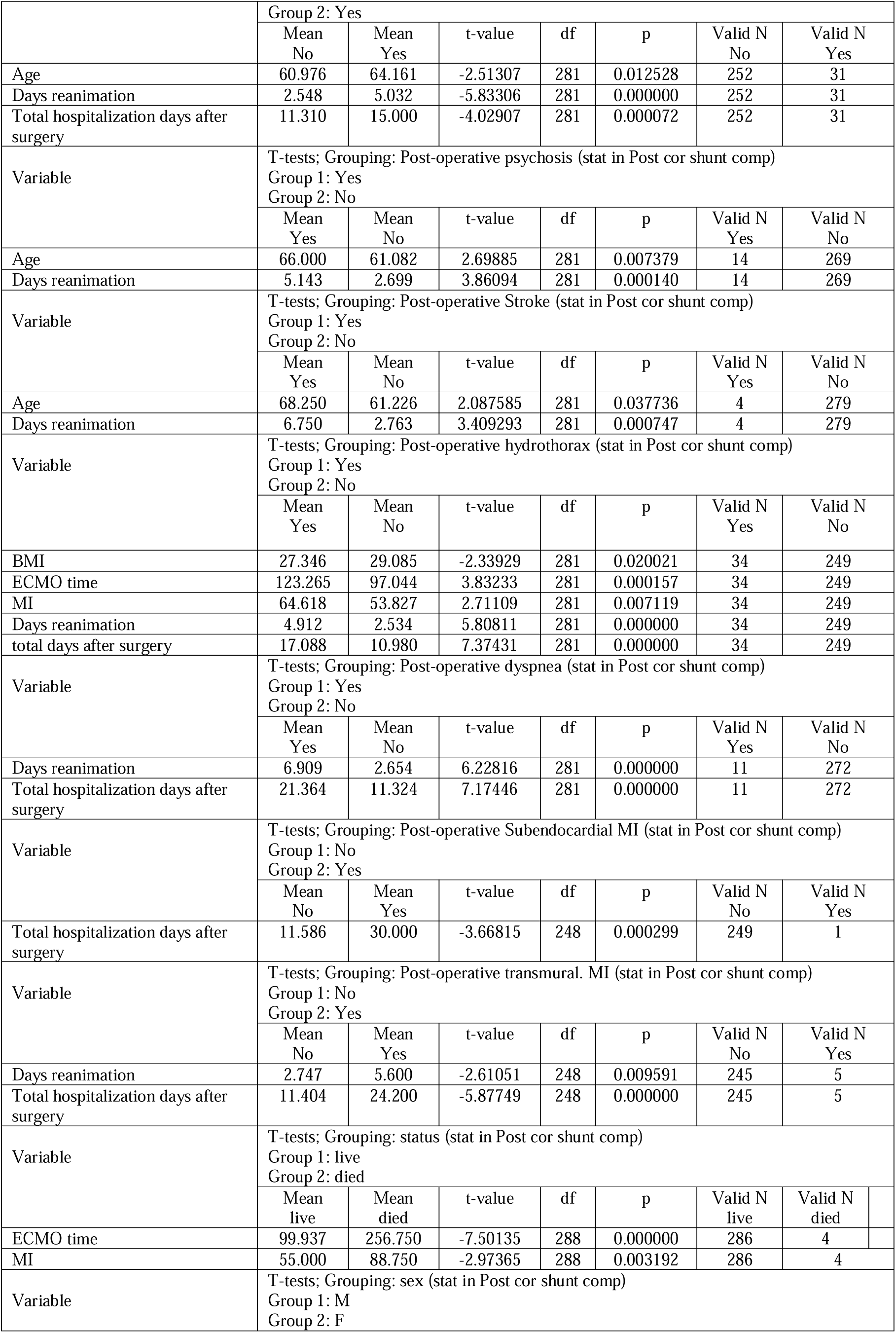
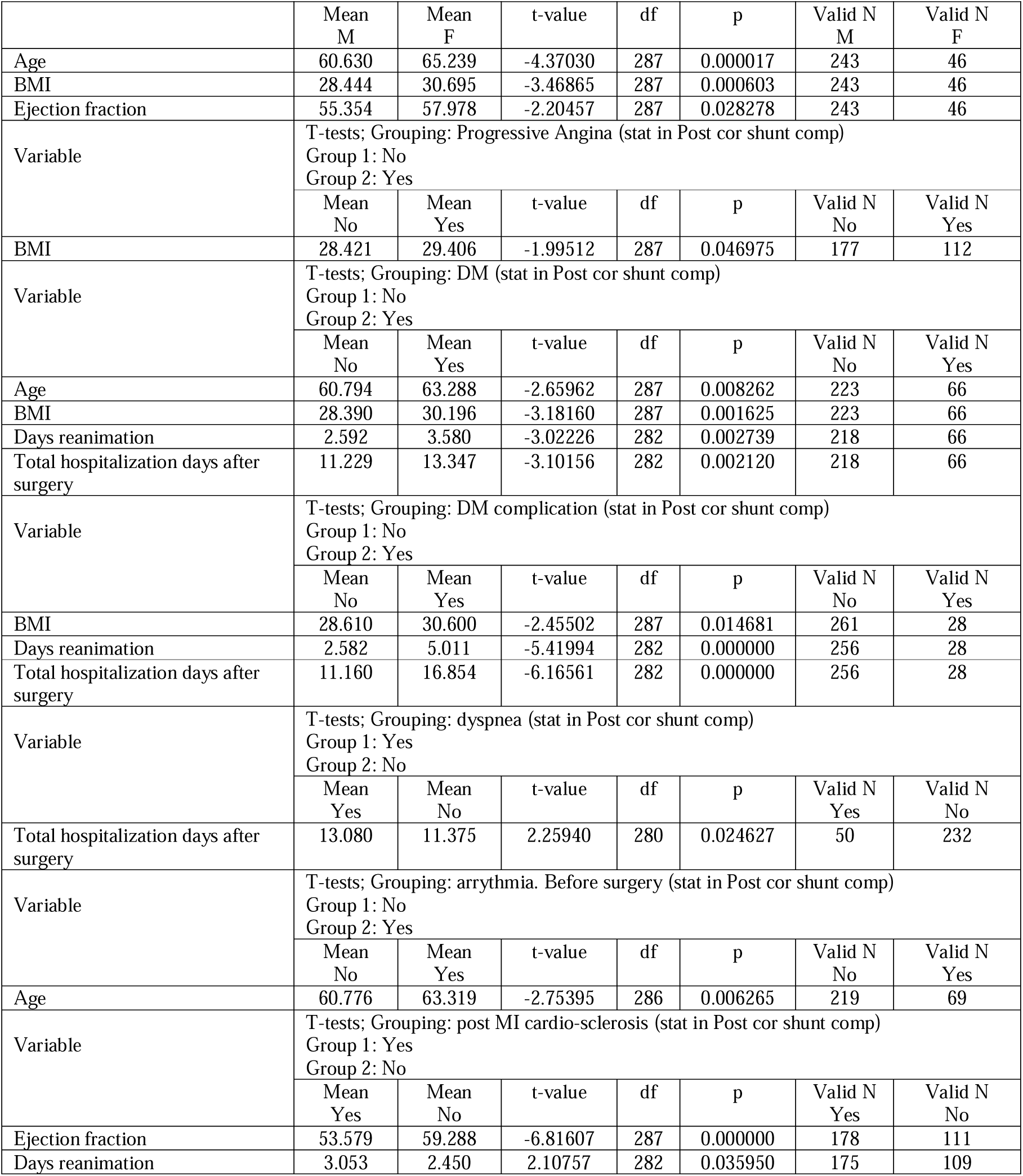
Comparation demonstration of various factors in the development of different complications in post CABG survivals.

In terms of post-operative complications. patients with coronary artery bypass graft (CABG) and plastic surgery repair of the left ventricle aneurysm had longer Aortic cross-clamp time. t-value -2.20413, p <0.028306. Additionally, patients with CABG have less ejection fraction, t-value 5.10667, p < 0.000001. Patients with post-CABG left ventricle (LV) aneurysm had less ejection fraction, t-value -2.01070, p <0.045292. Furthermore, patients with post-CABG LV aneurysm had a longer CPB time, t-value -5.58113, p < 0.000000. Furthermore, patients with LV aneurysm had longer Aortic cross-clamp time, t-value -4.72802, p < 0.000004.

Post-CABG arrythmia developed in elderly patients, t-value -2.51307, p<0.012528. Subsequently, after CABG arrythmia, the ICU and total hospitalisation days, t value - 5.83306, p< 0.000000; t-value -4.02907, p< 0.000072, respectively. Elderly people are at increased risk of psychosis after CABG surgery, t value 2.69885, p <0.007379. Postoperative psychosis significantly increases ICU hospitalization days, t-value 3.86094, p < 0.000140.

Postoperative stroke occurs more frequently in elderly people, t-value 2.087585, p < 0.037736. Subsequently, postoperative stroke increases the ICU hospitalization days, t-value 3.409293, p <0.000747. The incidence rate of complications in the first and second groups is shown in Figure 4.

**Figure 4:**
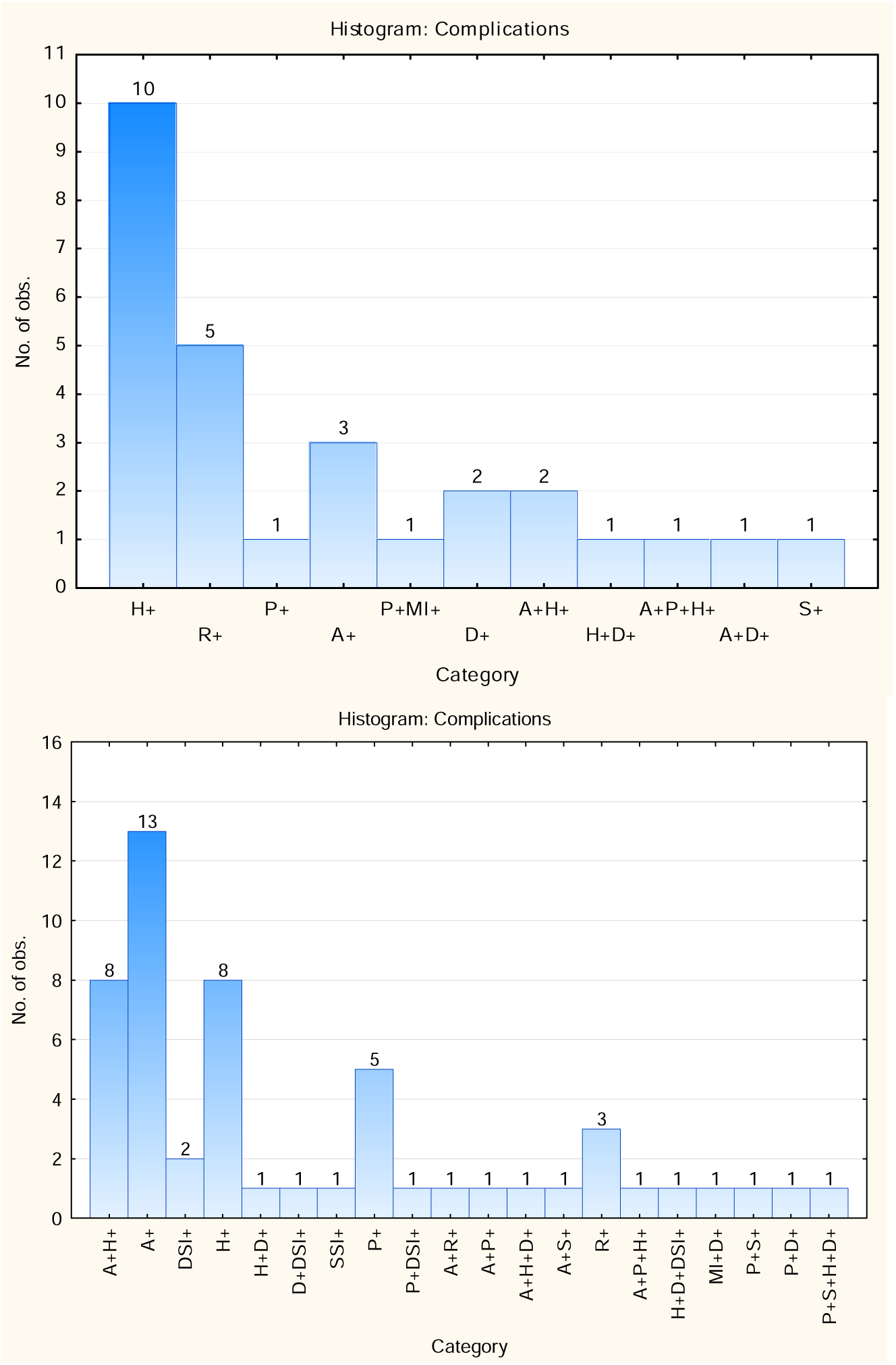
The incidence rate of complications in the first (<60 years) and second group (> 60 years), respectively. (H; hydrothorax, R; resternotomy, A; arrythmia, P; psychosis, MI; myocardial infarction, D; dyspnea, DSI; deep sternum wound infection, SSI; superficial sternum wound infection)

The number of ITAs used is less in the elderly, t-value 2.41992, p < 0.016145. In terms of correlation, we found a direct association between age and ICU / total hospitalisation days / number of complications, r= 0.189046, 0.141415, 0.138565; respectively. No statistical significance in the correlation between number of the complications and the number of the shunts as well as between the number of used internal thoracic arteries and number of complications.

## Discussion

In light of results, prolonged Aortic cross-clamp time is observed in coronary artery bypass graft (CABG) and plastic surgery repair of the left ventricle aneurysm and patients with preoperative LV aneurysm. The prolonged Aortic cross-clamp time results in the use of two internal thoracic arteries. The prolonged aortic cross-clamp time increases the risk of hydrothorax and the number of shunts, which subsequently increases the CPB time. Patients with low BMI are at high risk of hydrothorax. Furthermore, hydrothorax and the use of two internal thoracic arteries increase the time of the CPB. The coexistence of CABG and left ventricle aneurysm increases the time required for CPB.

The post CABG ICU hospitalization days is related with the presence of post-operative arrythmia, post-operative psychosis, post-operative stroke, post-operative hydrothorax, post-operative dyspnea, and superficial sternal wound infection. Total hospitalization days depend on the presence of arrythmia, which is commonly seen in elderly patients > 63 years old. Additionally, deep sternal wound infection increases total hospitalisation days and hospitalisation days. The ejection fraction and the days of hospitalisation in the ICU are closely associated with the total days of hospitalisation. Ejection fraction is age, sex, and presence of CABG and or CABG with LV aneurysm plastic repair dependent.

Age is a crucial factor in terms of postoperative complications. Where advancement in age increases the days of hospitalisation in the ICU, the total days of hospitalisation, and the number of complications. Elderly people are at high risk for stroke, psychosis, and arrythmia [4,7–14]. However, elderly people passed one internal thoracic artery CABG.

The number of complications is not associated with the death and alive status of patients or with the number of shunts. Furthermore, there were no statistical differences in the number of complications and the number of shunts and the number of internal thoracic arteries used. However, the number of complications depends on the existence of preoperative progressive angina and PMIMS. In addition, the number of complications is determined by age increase, CPB time, the Aortic cross-clamp time, the days of hospitalization in the ICU, and total hospitalization days.

The results of our study are consistent with those of other studies. Several studies reported that post-CABG complications include arrythmia, particularly atrial fibrillation [4,15–21]. However, the potential underlying pathophysiological mechanisms remain unclear and further elaboration is required. However, the potential pathophysiological pathway is multifactorial and involves autophagy dysfunction, upregulated sympathetic tone, mitochondrial dysfunction, inflammation, abnormal atrial conduction, and systemic inflammatory response indicated by elevated C-reactive protein and leukocytes [22–31]. Some studies showed that on-pump CABG surgery has higher risk of atrial fibrillation than off-pump [32–37].

## Conclusions

Hydrothorax is seen in a low BMI and a long Aortic cross-clamp time which leads to longer CPB time. Progressive angina and PMIMS increase the risk of postoperative complications, particularly hydrothorax. Elderly people undergoing CABG are at higher risk of psychosis, arrythmia, longer total and ICU hospitalization days, and stroke.

## Supporting information

Graphical abstract

## Data Availability

The data are applicable on reasonable request.

## List of abbreviations

CVD: Cardiovascular disease
COVID-19: Corona virus infectious disease-19
CABG: coronary artery bypass graft
CPB: cardiopulmonary bypass
BMI: body mass index
MI: myocardial infarction
DM: Diabetes mellites
ICU: intensive care unit
PMIMS: post-myocardial infarction myocardial sclerosis
LV: left ventricle
ITA: internal thoracic artery

## Declarations

1. Ethics approval and consent to participate: The study approved by the National Research Mordovia State University, Russia, from “Ethics Committee Requirement N8/2 from 30.06.2022”.
2. Consent for publication: Written informed consent was obtained from the participants for publication of study results and any accompanying images.
3. Availability of data and materials: applicable on reasonable request.
4. Competing interests: The authors declare that they have no competing interests regarding publication.
5. Funding’s: The work of **Basheer Abdullah Marzoog** was financed by the Ministry of Science and Higher Education of the Russian Federation within the framework of state support for the creation and development of World-Class Research Center ‘Digital biodesign and personalized healthcare’ № 075-15-2022-304.
6. Authors’ contributions: MB is the writer, researcher, collected and analyzed data, and revised the manuscript, EV collected the primary data from the hospital. All authors have read and approved the manuscript.
7. Acknowledgments: not applicable
8. Authors’ information: **Basheer Abdullah Marzoog**, Research Center «Digital Biodesign and Personalized Healthcare», I.M. Sechenov First Moscow State Medical University (Sechenov University), 119991 Moscow, Russia; postal address: Russia, Moscow, 8-2 Trubetskaya street, 119991. (, +79969602820). ORCID: 0000-0001-5507-2413. ORCID: 0000-0001-5507-2413. Scopus ID: 57486338800. Ekaterina Vanichkina, National Research Ogarev Mordovia State University. ORCID: 0009-0006-3015-2306
9. The paper has not been submitted elsewhere

