## Supplementary figures and images for "Incidence Rate of Post Coronary Artery Shunt Complications; Age Dependent!"

### Graphical abstract

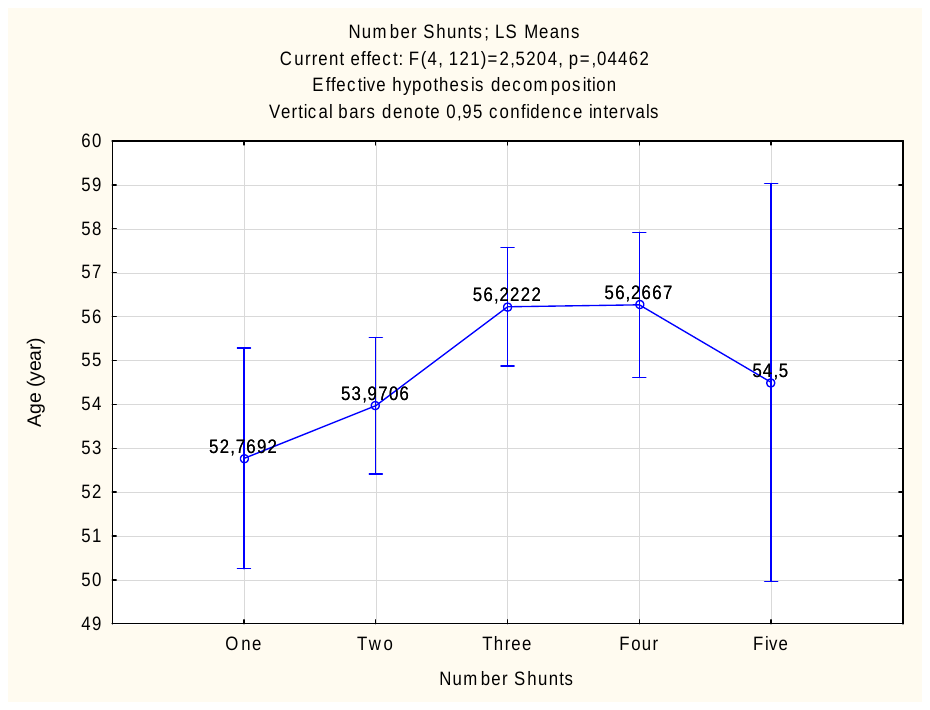

Figure 4: With age, the number of shunts increases in the first group < 60 years.
